# MONITORING OF RESPIRATORY VIRUS COINFECTION IN SOUTHERN BRAZIL DURING COVID-19 PANDEMIC

**DOI:** 10.1101/2024.04.09.24305542

**Authors:** Dayane Azevedo Padilha, Fernando Hartmann Barazzetti, Marcos André Schörner, Vilmar Benetti Filho, Eric Kazuo Kawagoe, Doris Sobral Marques Souza, Maria Luiza Bazzo, Glauber Wagner, Gislaine Fongaro

## Abstract

Since December 2019, the COVID-19 pandemic caused by SARS-CoV-2 has reached approximately 769 million people, leading to more than 7 million deaths worldwide. Faced with the possible presence of other respiratory pathogens that could co-infect and modify the clinical response of patients detected for SARS-CoV-2, some researchers have explored this line of investigation. The relationship between these co-infections remains unclear, leading to a need to deepen our knowledge about interactions among pathogens, and between pathogens and the host. Thus, the present study employed RT-qPCR to assess the presence of Human Adenovirus (HAdV), Influenza A (Flu A), Influenza B (Flu B), Human Metapneumovirus (HMPV), Respiratory Syncytial Virus (RSV), Human Rhinovirus (HRV), and Parainfluenza Virus (PIV). A total of 187 nasopharyngeal samples from adult patients exhibiting respiratory symptoms were collected between February 2021 and November 2022 at the University Hospital Polydoro Ernani de Sao Thiago in Florianopolis, SC, Brazil. Our findings revealed that 25.16% of samples tested positive for non-SARS-CoV-2 respiratory viruses (29.8% - HRV, 5.3% - PIV, 4.3%-RSV, and 1.1% - HMPV). From the 74.84% of SARS-CoV-2 positive patients, the presence of co-infection was observed in 9,7% of patients, with 7.5% being HRV, 1.1% HAdV and 1.1% Flu A. Since co-infections can potentially alter patient prognoses and impact local epidemiological dynamics, this study highlights the significance of ongoing monitoring and epidemiological assessment through genomic surveillance of other clinically relevant respiratory pathogens.

## 1 INTRODUCTION

In August 2023, the pandemic of COVID-19, caused by SARS-CoV-2, reached approximately 769 million cases and almost 7 million deaths worldwide, while in Brazil, the number of cases is approaching 37 million with approximately 700,000 deaths. These data reinforce the clinical and epidemiological importance of this pandemic that lasted more than 3 years 1.

Early reports from China said that co-infection of SARS-CoV-2 with other respiratory viruses is rare. However, studies have shown that the presence of co-infection between SARS-CoV-2 and other respiratory viruses seems to aggravate lung disease in such a way as to increase the need for mechanical ventilation (when co-infected with influenza, for example), in addition to impacting the circulation of seasonal viral infections and patient morbidity in cases where there is co-infection of SARS-CoV-2 with Respiratory Syncytial Virus (RSV), Human Adenovirus (HAdV), Human Rhinovirus (HRV) and human Metapneumovirus (HMPV) 2, 3.

The co-infection of SARS-CoV-2 and influenza is the most reported in the literature. In addition, the two viruses share similarities in terms of respiratory symptoms. It is believed that at the beginning of the pandemic, several cases may have been misdiagnosed, as at that time there were no well-established diagnostic tests for SARS-CoV-2 4.

During the first cases of severe acute respiratory infections, at a time in which the pathological agent had not been identified, molecular biology techniques for detecting pathogens through respiratory panels were decisive and paved the way for the sequencing and classification of SARS-CoV-2. At the beginning of the SARS-CoV-2 pandemic, the number of studies of co-infection with other respiratory pathogens was very low. Even today, after the WHO declared the end of the pandemic, knowledge about the influences of co-infections of SARS-CoV-2 with other respiratory viruses is unclear with regards to host-pathogen interactions, patterns of infection and transmissibility, and the clinical outcome of patients 5.

Studies have demonstrated the importance of identifying pathogens that cause acute pneumonia even before COVID-19, given the pulmonary predominance of Influenza A (Flu A) and Influenza B (Flu B), HRV, RSV, HMPV, and Parainfluenza Virus (PIV), which had their epidemiological patterns modified with the insertion of SARS-CoV-2 into the population 6.

The present study evaluated 187 nasopharyngeal samples from patients with non-serious respiratory symptoms, admitted to a screening unit for the diagnosis of COVID-19, to determine co-infections by HAdV; Flu A; Flu B; HMPV; RSV; HRV, an**d PIV**. Thus, this study aims to report the profile of co-infections of respiratory viruses and SARS-CoV-2 from February 2021 to November 2022 in the state of Santa Catarina, Brazil.

## 2 MATERIALS AND METHODS

### 2.1 SAMPLES PROCESSING

A total of 187 nasopharyngeal samples from adults (men and women), with non-serious respiratory symptoms, were randomly collected between February 2021 and November 2022 – of which 88 were obtained from healthcare professionals – with 93 positive and 94 negative diagnoses for SARS-CoV-2. Samples were collected using a nasopharyngeal swab in an appropriate transport medium, as recommended by health agencies, for the diagnosis of SARS-CoV-2.

### 2.2 RNA PURIFICATION AND PCR

Samples were aliquoted and stored in cryotubes at −80°C. Viral genetic material was extracted using QIAmp Viral RNA Mini Kit (Qiagen, USA). Detection of SARS-CoV-2 was performed using the Allplex™ 2019-nCoV Assay Kit or Allplex™ SARS-CoV-2 Assay Kit (Seegene, Korea). Negative and positive samples for SARS-CoV-2 from symptomatic patients were tested in the Allplex™ RV Essential Assay (Seegene, Korea), multiplex qPCR that detects seven viruses: HAdV; Flu A; Flu B; HMPV; RSV; HRV, and PIV. Amplification was performed according to the manufacturer’s instructions on the CFX96™ Real-time PCR System thermal cycler (Bio-Rad®) and the results were visualized using the Seegene View software.

### 2.3 STATISTICAL ANALYSIS

During this study, 187 nasopharyngeal samples from adult patients with respiratory symptoms were collected between February 2021 and November 2022 at the Hospital Universitário Polydoro Ernani de São Thiago in Florianópolis, SC, Brazil. After PCR testing for the viruses HAdV, Flu A, Flu B, HMPV, RSV, HRV, and PIV, Fisher’s exact test was used to examine the association between SARS-CoV-2 and other viral infections.

## 3 RESULTS

From February 2021 to November 2022, 187 samples were randomly selected to determine co-infection of SARS-CoV-2 with HAdV, Flu A, Flu B, HMPV, RSV, HRV, and PIV. None of the 187 samples were positive for Flu B. Between February 2021 and November 2022, one test was positive for HAdV, one for Flu A, one for HMPV, four for RSV, 35 HRV, and five for PIV.

Absolute and relative frequencies for viral infections are shown in Figure 1. HRV was the most frequent infection for patients with negative qPCR results for SARS-CoV-2 (Figure 2) and showed co-infections with PIV, HMPV, and RSV (Figure 1). SARS-CoV-2-positive patients had more co-infections with HRV than the other tested viruses (Figure 2). Also, Flu A and HAdV were only detected in SARS-CoV-2 positive patients and this result indicates a relationship between SARS-CoV-2 and other viral infections caused by Influenza A and HAdV (Fisher’s exact test p-value = 0.000001327).

**Figure 1.**
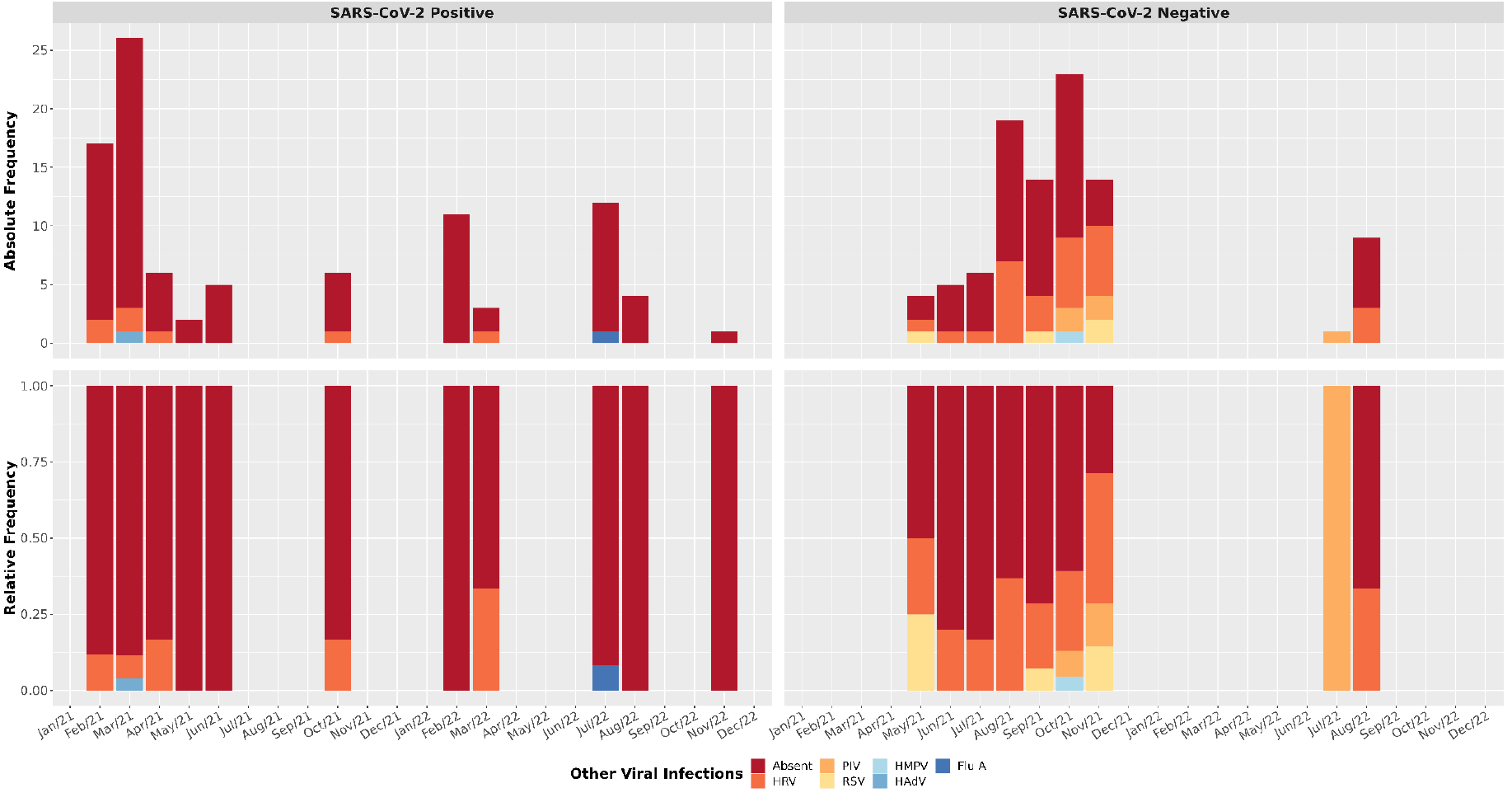
Viral profile in positive and negative SARS-CoV-2 samples. Absolute and relative frequency for respiratory viral panel tests. Human Adenovirus (HAdV); Influenza A (Flu A); Human Metapneumovirus (HMPV); Respiratory Syncytial Virus (RSV); Human Rhinovirus (HRV) and Parainfluenza Virus (PIV).

**Figure 2.**
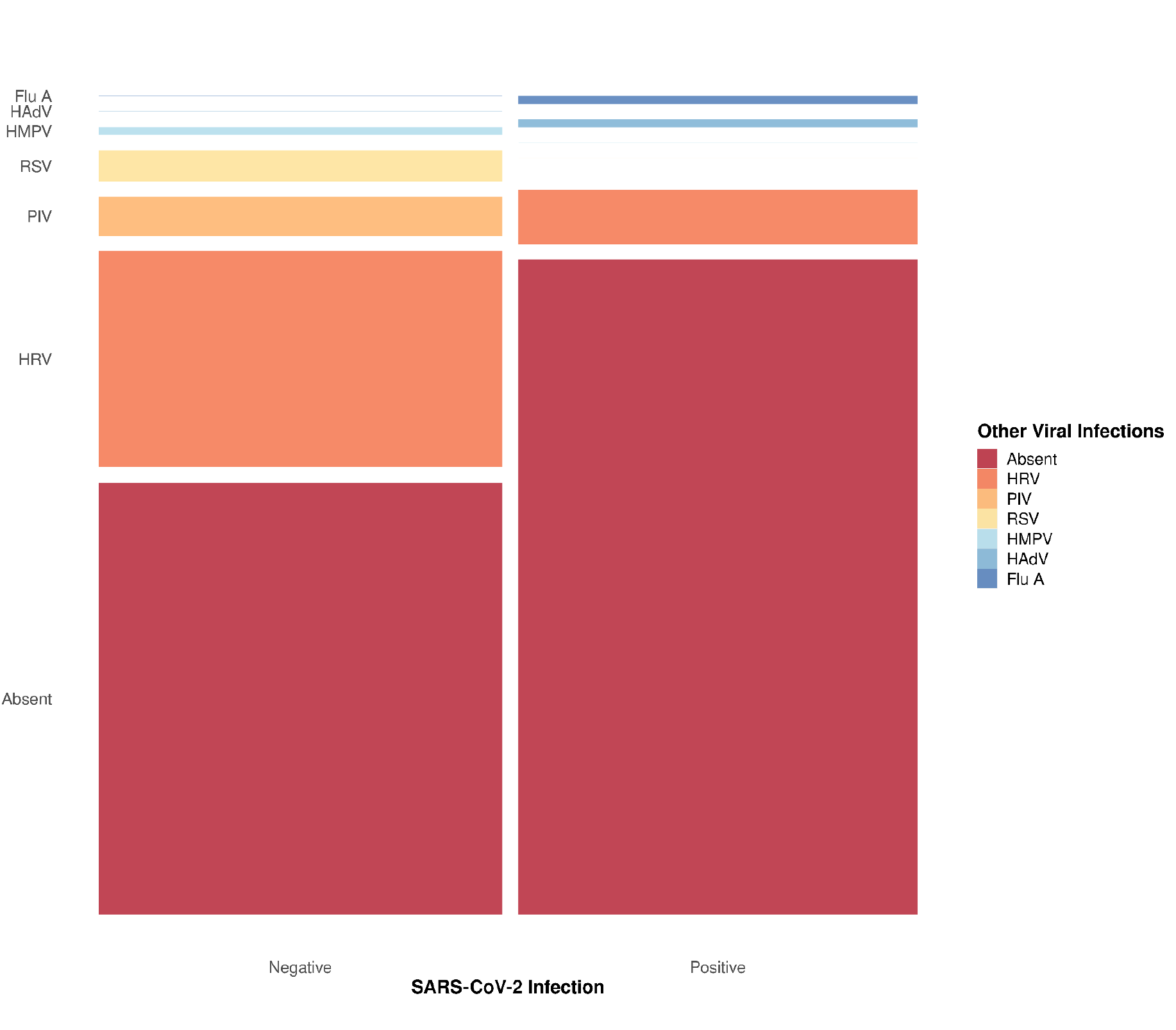
Summary of viral infection related to SARS-CoV-2 diagnostic. The respiratory viral panel tests included Human Adenovírus (HAdV), Influenza A (Flu A), Human Metapneumovirus (HMPV), Respiratory Syncytial Virus (RSV), Human Rhinovirus (HRV), and Parainfluenza Virus (PIV). The thinnest lines represent zero counts and all viruses are ordered in the same sequence in both columns.

## 4 DISCUSSION

Even after the official WHO declaration 1 on the end of the public health emergency of international concern related to COVID-19, scientific data still needs to be explored in search of answers that can elucidate the variations in the clinical presentation among patients and in the epidemiological aspects of SARS-CoV-2. One of these explorations involves the importance of co-infection with other respiratory pathogens 1.

Respiratory viruses such as influenza (Flu A and Flu B) and RSV were already known in the pre-pandemic periods for affecting the population, generating epidemic outbreaks with cardiorespiratory impairment, which could lead to morbidity in the infected population 7,8. Studies have shown that even early in the pandemic, diagnosis of other respiratory pathogens helped identify the virus that later became known as SARS-CoV-2. Some studies have reported the clinical importance of patients co-infected with respiratory viruses combined with SARS-CoV-2, identifying an increased need for mechanical ventilation after co-infection 3,9.

The epidemiological data show a clear change in the detection of seasonal viruses such as influenza, which seems not to have been identified between the years 2020 and 2022, the period with the highest number of cases and deaths caused by COVID-19 10. It is not clear if this lower detection of seasonal respiratory viruses is due to the reduction in the prevalence of pathogens other than SARS-CoV-2 owing to interactions and competition between pathogens, to a possible underreporting resulting from the overload of health professionals involved in the pandemic, or both 11.

Of the total of 187 samples from adult patients (men and women) with non-serious respiratory symptoms, one test was positive for HAdV, one for Flu A, one for HMPV, four for RSV, 35 for HRV, and five for PIV. Our data corroborates Kim and collaborators who, in 2020, had already demonstrated higher report rates of HRV and RSV infections in patients with respiratory symptoms but undetected for SARS-CoV-2 11,12.

It is important to highlight that, during their study, Kim and collaborators 12 also evaluated the presence of enterovirus, Chlamydia pneumoniae and Mycoplasma pneumoniae, unlike our study, in which such infections may have been underreported, and which have clinical importance and may worsen the prognosis of patients infected with SARS-CoV-2. In addition, the number of cases of COVID-19 in our study was much higher 12,13.

Another study 14 evaluated, between November 2021 and February 2022, the presence of co-infections among patients detected with SARS-CoV-2 in combination with Flu A (65%), enterovirus-rhinovirus (20%), HAdV (2%), RSV (2%), HMPV (2%), Human parainfluenza virus type 3 (2%), and Human coronavirus (7%). Unlike our study, in which patients detected for SARS-CoV-2 had a higher prevalence of co-infection with HRV, Eldesouki et al. 14 had the highest prevalence of co-infection between SARS-CoV-2 and Flu A. On the other hand, we must consider that the studies were developed with different sample sizes (41 patients versus 187), in addition to differences in population characteristics 14.

In Brazil, since 2021, data from the Board of Epidemiological Surveillance of the State of Santa Catarina - DIVE (official website for monitoring infectious diseases) have presented RSV, HRV, HAdV, bocavirus, in addition to the unidentified ones, as the main agents causing acute respiratory syndrome after SARS-CoV-2. These data are partially different from our results, showing HRV as the main agent detected both in patients detected and undetected for SARS-CoV-2 15.

Severe Acute Respiratory Syndrome (SARS) is a more aggressive infection that can lead to hospitalizations and death. In the surveillance carried out in this work, the largest number of infections reported, except for SARS-CoV-2, is by HRV. This discrepancy in the data can be explained in part by the sampling of this work, which did not cover severe cases, but only classic flu symptoms, a profile different from that monitored by public agencies. Interestingly, although our results demonstrate only one sample of co-infection between SARS-CoV-2 and Flu A, epidemiological data from DIVE 16,17 reported 72 deaths in the state of Santa Catarina due to SARS caused by Flu A. We hypothesize that this difference is due to the population group of the study, which were symptomatic adults without enough clinical severity to justify hospitalization characteristics of SARS.

## 5 CONCLUSIONS

Between February 2021 and November 2022, respiratory viruses were diagnosed in 187 adult patients of both sexes, with characteristic symptoms of respiratory infection. During this study, HRV, PIV, RSV, and MPV were detected in patients not detected for SARS-CoV-2, in addition to the detection of HAdV, Flu A, and HRV co-infecting patients detected for SARS-CoV-2. These data demonstrate the importance of the monitoring of contagious infectious pathogens of clinical and epidemiological importance, in addition to SARS-CoV-2, aiming to identify the population epidemiological pattern to improve the tools for preventing transmissibility and making therapeutic approaches as individualized as possible.

## Data Availability

All data produced in the present work are contained in the manuscript

## DISCLAIMER

The opinions expressed by authors contributing to this journal do not necessarily reflect the opinions of the Federal University of Santa Catarina, Brazil, or the institutions with which the authors are affiliated. The funders had no role in the study design, data analysis, or the decision to publish.

## INSTITUTIONAL REVIEW BOARD STATEMENT

The study was conducted in accordance with the Declaration of Helsinki and approved by the Ethics Committee in Research Humans Beings of Federal University of Santa Catarina (Assent number: 4.035.636, CAAE: 31521920.8.0000.0121, 19 May 2020).

## ACKNOWLEDGMENTS

This work is dedicated to all SC citizens who suffered or passed away due to COVID-19. We are indebted to Hospital Universitário/HU/EBSERH at the Federal University of Santa Catarina (Florianópolis, SC, Brazil) for granting access to interview patients and perform sample collection.

